# Effectiveness and safety of a New Standardised TrEatment Protocol based on initial/early therapy with single-pill combinations of blood pressure-lowering drugs for improving blood pressure control (NewSTeP): Rationale, design of a randomized clinical trial and baseline characteristics of the trial participants

**DOI:** 10.64898/2026.09.10.26362712

**Authors:** Rupasvi Dhurjati, Rashmi Pant, Amit Kumar, Anshika Mittal, Krishnaiah Chappidi, Gautam Satheesh, Parul Puri, Sasi Kumar Tiruttani, Shwetha Rajaram, Aneesh Basheer, Prabhdeep Kaur, Andrew Moran, Andres Rosende, Poongothai Subramani, Sandeep Mahajan, Nupur Lalvani, Sandeep Bansal, Dorairaj Prabhakaran, Jayagopal Pathiyil Balagopalan, Stephen Jan, Mark D. Huffman, Vivekanand Jha, Abdul Salam

## Abstract

**Background:** Hypertension is the leading modifiable risk factor for cardiovascular disease (CVD). Despite availability of blood pressure (BP)-lowering therapies, BP control rates remain suboptimal. Reluctance to initiate treatment with combination therapy and delayed treatment intensification contribute to poor control. Early use of single-pill combinations (SPCs) within standardised treatment protocols (STPs) may improve outcomes, but evidence from India is limited.

**Objective:** To evaluate the effectiveness and safety of a new STP incorporating initial or early use of SPCs compared with usual care among adults with hypertension.

**Methods:** The New Standardised TrEatment Protocol (NewSTeP) trial is a pragmatic, multicentre, randomized, parallel-group, open-label trial conducted in tertiary healthcare centres in India. Adults with hypertension who are untreated or receiving one BP-lowering drug and require treatment initiation or intensification (n = 300) will be randomized 1:1 to either a new STP incorporating early SPC therapy or usual care. Participants will be followed for 6 months. The primary effectiveness outcome is change in home-measured mean systolic BP from randomization to month 6. The primary safety outcome is discontinuation of trial drug due to adverse events. Secondary outcomes include proportion of participants achieving BP control, change in diastolic BP, incidence of adverse events of special interest, treatment adherence, treatment modification, and time to BP control. A process evaluation will assess reach, implementation, acceptability, and sustainability, while an economic evaluation will assess cost-effectiveness.

**Results:** Between October 2025 and March 2026, 300 participants were randomized across nine sites (150 to New STP; 150 usual care) from 360 screened individuals. Final follow-up is expected in September 2026, with results anticipated in October 2026.

**Conclusions:** The NewSTeP trial will provide evidence on the effectiveness, safety, implementation, and cost-effectiveness of an SPC-based STP for hypertension management in India and inform future hypertension control strategies and policies.

**Trial Registration:** The trial is registered with the Clinical Trials Registry–India (CTRI/2025/07/091649).

## Introduction

Cardiovascular diseases (CVDs) are the leading cause of mortality in India, accounting for more than one-quarter of all deaths.[1] Hypertension remains the single most important modifiable risk factor for CVD and affects approximately 30% of the adult population. Although effective blood pressure (BP)-lowering treatments are widely available, BP control rates remain suboptimal, with national estimates suggesting that only one-third of adults with hypertension are aware of their condition, 15% are on treatment, and only about 13% achieve recommended BP targets.[2–6] Despite recent efforts to strengthen hypertension care, including large-scale programmes such as the Indian Hypertension Control Initiative (IHCI), BP control rates remain below 50%. [7] These persistently low rates of BP control reflect important gaps between evidence and routine clinical practice.

These gaps arise from a combination of patient-, provider-, and health system-level barriers. At the provider level, treatment is often initiated with a single BP-lowering drug (monotherapy) within a four- or five-step treatment algorithm, and subsequent therapeutic inertia may delay treatment intensification, prolonging exposure to uncontrolled BP [8, 9]. Most of the patients treated by existing protocols will eventually require two or more BP-lowering drugs to achieve target BP.

In response to these challenges, international hypertension guidelines increasingly recommend the use of combination BP-lowering therapy as initial treatment for most patients with hypertension.[10–12] Single pill combinations (SPCs), which combine two or more antihypertensive drugs in a single pill, simplify treatment regimens, improve adherence and can achieve greater BP reduction than monotherapy without substantially increasing adverse effects.[13, 14] Meta-analyses and randomized controlled trials have consistently demonstrated superior BP control with combination therapy compared with conventional monotherapy-based treatment strategies.[15–17]

Standardised treatment protocols (STPs) provide another promising strategy to improve hypertension management. STPs offer structured guidance for treatment initiation, intensification and follow-up, reducing unwarranted variation in care and facilitating implementation of evidence-based treatment pathways. The World Health Organization (WHO) HEARTS technical package and Pan-American Health Organization (PAHO) identify the adoption of a context-appropriate STP as a key component of successful hypertension control programmes.[18–20]

Although evidence supporting early use of SPCs is growing, data from India evaluating the effectiveness and safety of SPC-based hypertension protocols remain limited. The New Standardised TrEatment Protocol (NewSTeP) trial was therefore designed to develop and evaluate a contextualised hypertension STP incorporating initial/early use of SPCs in Indian healthcare settings. Through a combination of formative research, a pragmatic randomized controlled trial, process evaluation and economic evaluation, the trial aims to generate evidence to inform future hypertension control programmes and policy implementation for improving BP control in India. This protocol describes the methodology of the NewSTeP RCT, process evaluation and economic evaluation.

## Objectives

To investigate, among adults with high BP who are either untreated or receiving one BP-lowering drug, the effectiveness and safety of a new SPC based STP (New STP) based on initial/early therapy with single-pill combinations of BP-lowering drugs, compared with usual care.

## Methods

### Design

The NewSTeP trial is a pragmatic, randomized, parallel-group, open-label trial comparing a new STP with usual care over a 6-month treatment and follow-up period. The trial schema is shown in Figure 1. The trial is registered with the Clinical Trials Registry–India (CTRI/2025/07/091649).

**Figure 1:**
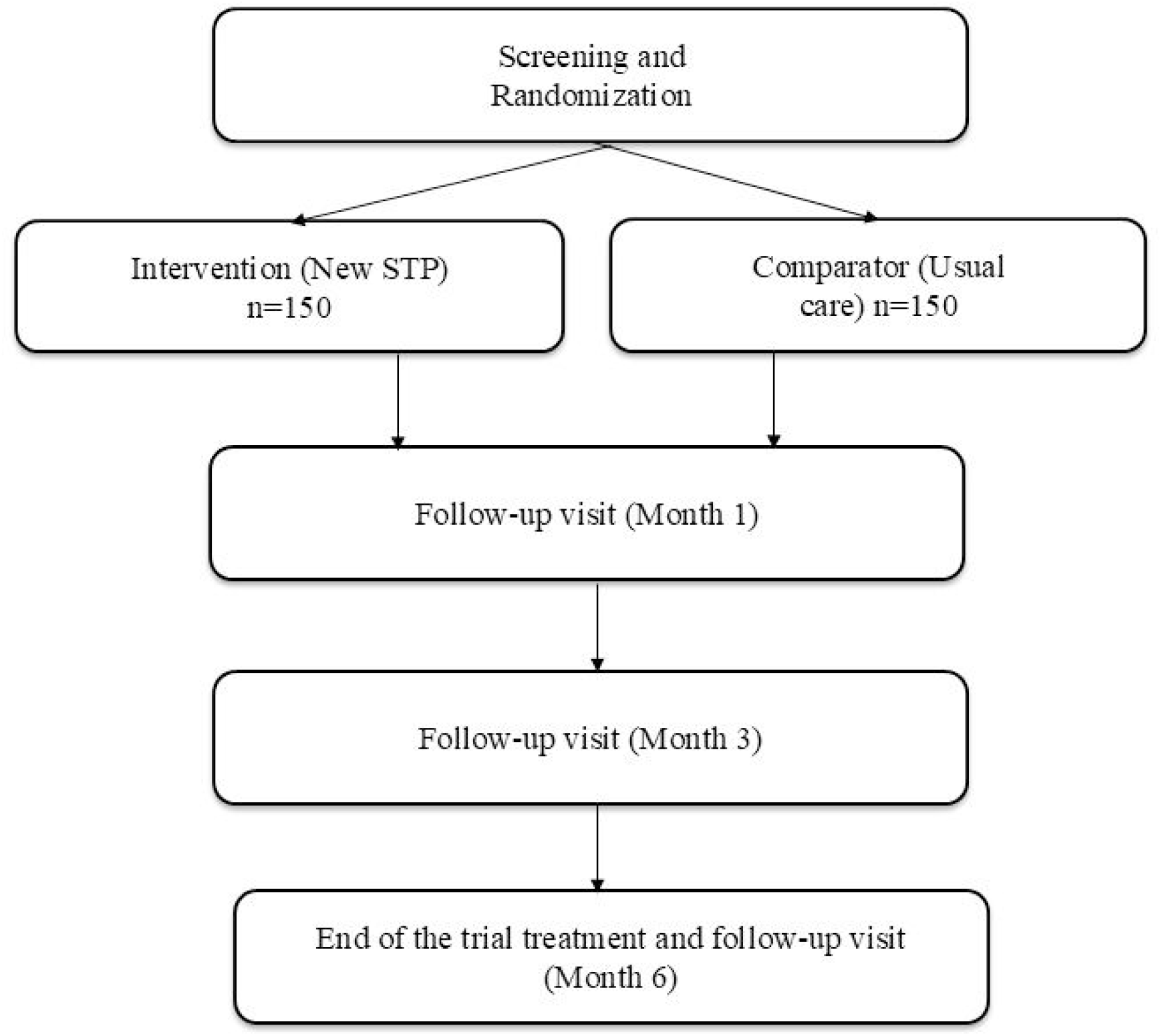
NewSTeP trial schema. Note: For participants in the intervention arm, the additional follow-up visits are conducted as per the new hypertension STR and for participants in the control arm, as per usual care.

### Settings

The participants will be recruited from the outpatient departments of purposively selected healthcare centres providing hypertension care in India. The sites will be selected based on their capacity to recruit patients with hypertension as per the recruitment plan, conduct the trial procedures and support trial follow-up, while also taking into account the diversity in geographic location of the site and patient characteristics.

### Randomization

Participants will be centrally assigned in a 1:1 ratio to the new STP or usual care arm using a computer-generated block randomization sequence, prepared by an independent statistician using variable block sizes (2 and 4), stratified by trial site and baseline BP-lowering treatment status (0 or 1 drug), through the electronic case report form (eCRF), ensuring allocation concealment until participant assignment. Blinding of participants and site staff is not feasible given the open-label nature of the intervention and comparator.

### Participant eligibility

Adults with hypertension, who are either untreated or treated with one BP-lowering drug, and who could appropriately be treated with the new STP or usual care for hypertension and have no contraindication to trial drugs or treatment will be enrolled. Detailed inclusion and exclusion criteria are reported in Table 1.

**Table 1:** Participant eligibility criteria.

| Inclusion Criteria |
| --- |
| *At screening visit*<br>Adults (age 18- 79 years)<br>Have diagnosis of hypertension<br>Documented previous diagnosis of hypertension and/or currently using BP-lowering drugs for hypertension<br>Clinic attended automated seated mean systolic blood pressure (SBP) $\geq 140$ mmHg and/or diastolic blood pressure (DBP) $\geq 90$ mmHg on screening visit day and<br>At least one documented instance of $\geq 140$ mmHg and/or DBP $\geq 90$ mmHg in the past<br>Untreated for hypertension or on one BP-lowering drug for $\geq 2$ weeks.<br>Needs initiation or intensification of BP-lowering drug(s) as per the investigator’s judgement based on BP at this and/or previous visits<br>Willingness to use a home BP measurement device and measure BP at home for 6 months<br>Provided signed informed consent to participate in the trial. |
| *At randomization visit*<br>1. Untreated or on one BP-lowering drug for $\geq 2$ weeks<br>2. Clinic attended automated seated mean SBP (average of last 2 measurements): 140-179 mmHg and/or DBP 90-109 mmHg<br>3. Willingness to use home BP measurement device and measure BP at home for 6 months<br>4. Needs initiation or intensification of BP-lowering drug(s) as per the investigator's judgement based on BP at this and/or previous visits. |
| **Exclusion Criteria** |
| *At screening visit*<br>1. Receiving 2 or more BP-lowering drugs (SPCs containing two BP-lowering drugs should be considered as 2- BP lowering drugs)<br>2. Receiving any BP lowering drugs for primary indications other than hypertension (e.g., migraine, benign prostate hyperplasia, heart failure)<br>3. Current/history of secondary hypertension, cardiovascular disease, including coronary heart disease, angina, myocardial infarction, acute coronary syndrome, congestive heart failure, atrial fibrillation, stroke or transient ischemic attack<br>4. Known current/history of end-stage renal disease or anuria or current estimated glomerular filtration rate (eGFR) $< 60$ ml/min/1.73 m<sup>2</sup>.<br>5. Women who are pregnant or had a positive pregnancy test or unwilling to take a pregnancy test before randomization and/ or during the trial, breastfeeding, or of childbearing potential and not using effective contraception during the trial period.<br>6. Contraindication, including hypersensitivity (e.g., anaphylaxis or angioedema) to any of the trial drugs or procedures<br>7. Participation in any investigational drug and/or device trial within the 30 days prior to randomization<br>8. Current concomitant illness or physical impairment or mental condition or abnormal laboratory value, which in the judgment of the investigator could interfere with the effective conduct of the trial or constitutes a significant risk to the participants' safety or well-being<br>*At randomization visit*<br>1. SBP $\geq 180$ or DBP $\geq 110$ mmHg<br>2. Fulfilling any of the exclusion criteria mentioned for the screening visit, when verified again |

### Interventions

Eligible participants will be randomized to either the new STP or usual care. Participants allocated to the new STP (Figure 2) will be initiated on treatment according to their average clinic BP and pre-randomization BP-lowering drug status. Those not receiving BP-lowering drug will be commenced on Step 1 treatment with one tablet of a dual SPC containing telmisartan 40 mg and amlodipine 5 mg, taken once daily. Participants already receiving one BP-lowering drug will be commenced on Step 2 treatment if their clinic SBP was ≥150 mmHg and/or DBP was ≥95 mmHg; otherwise, they will be commenced on Step 1 treatment.

**Figure 2:**
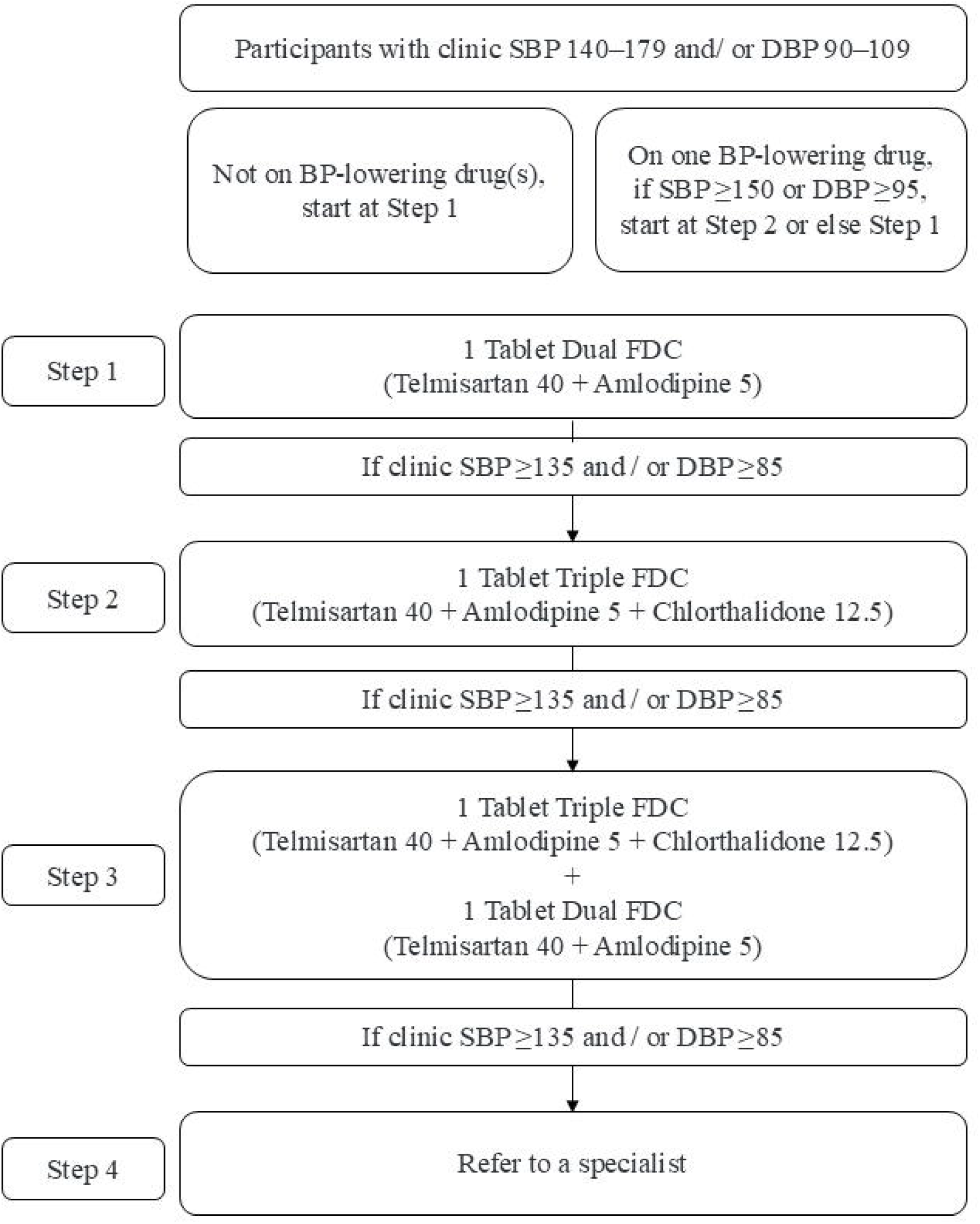
New Standardised Treatment Protocol.

Treatment will be sequentially intensified at scheduled follow-up visits if clinic BP remains ≥135/85 mmHg. At Step 2, participants will receive one tablet of a triple SPC containing telmisartan 40 mg, amlodipine 5 mg, and chlorthalidone 12.5 mg once daily. At Step 3, treatment may be escalated to one tablet of the triple SPC plus one tablet of the dual SPC (telmisartan 40 mg and amlodipine 5 mg), taken once daily. Participants whose BP remains above target despite Step 3 treatment will progress to Step 4 and will be referred to a hypertension specialist.

All trial drugs will be taken once daily, preferably at the same time each day. Down-titration will be considered in cases of hypotension (clinic or home SBP <90 mmHg or DBP ≤60 mmHg). Where elevated clinic BP is accompanied by poor adherence or persistently low home BP, treatment intensification could be delayed at the discretion of the treating clinician.

Participants randomized to usual care will receive antihypertensive treatment at the discretion of the treating physician.

### Study Visits and Procedures

Participants will be assessed for eligibility at the screening visit, during which baseline demographic and clinical information will be collected. Eligible participants will be randomized and initiated on their allocated treatment strategy. Screening and randomization may be conducted on the same day provided that protocol-required laboratory investigations performed within one month before screening are available and eligibility is confirmed.

Participants in the intervention arm will attend a total of six visits, while participants in the comparator arm will attend five visits. Participants in the intervention arm will be required to attend an additional visit at week 2 to assess treatment tolerance, drug adherence, and early BP response. Drug adjustments will be made at this visit, when indicated, in accordance with the trial protocol. All participants will attend follow-up visits at month 1, month 3, and month 6 following randomization. The month 6 visit will serve as the final follow-up visit (end of randomized period). (Figure 1)

During the trial, BP and pulse will be measured at all scheduled visits by trained site staff using a standardized clinic BP measurement procedure, with participants seated and BP recorded in triplicate. At the randomization visit, participants will be provided with a validated home BP monitoring device and a BP diary and will receive training on their use. To establish baseline home BP, participants in both the arms will be instructed to measure their BP in triplicate on the evening of the randomization day and again in triplicate on the following morning before taking the first dose of trial drug. Participants will also record their home BP in the morning and evening on four consecutive days immediately prior to the study visit and on a single set day of the participants’ preference in other weeks. The site staff will call the participants by phone and/or SMS at least once a week to check if the participants have taken their home BP measurements and recorded the values in the diary.

Home BP records and/or device data will be reviewed throughout the follow-up period, and additional training on home BP monitoring procedures will be provided when required. Physical examinations, clinic BP and pulse measurements, assessment of concomitant drugs, and evaluation of adverse events (AEs), adverse events of special interest (AESIs), and serious adverse events (SAEs) will be conducted according to the trial visit schedule. (Supplementary table 1)

Laboratory investigations will be performed during screening to determine eligibility and establish baseline safety assessments. Screening laboratory evaluations will include glucose (fasting or random), complete blood count, lipid profile, electrolytes, serum creatinine with estimated glomerular filtration rate (eGFR), thyroid-stimulating hormone (TSH), urinalysis/complete urine examination, pregnancy testing where applicable, and electrocardiography. A pregnancy test will be repeated at randomization if required. Electrolytes and creatinine with eGFR will be reassessed at months 3 and 6 to monitor participant safety. Additionally, glucose testing and lipid profile assessment will be repeated at month 6.

Lifestyle counselling will be provided at all the scheduled follow-up visits. The trial drugs will be dispensed at randomization, week 2, month 1, and month 3 visits. Returned drugs will be reviewed and collected at week 2, months 1, 3, and 6 to assess adherence. Healthcare utilisation and cost data will be collected at months 1, 3, and 6.

Unscheduled visits and additional follow-up visits may also be performed when clinically indicated. During such visits, BP, pulse rate, drug changes, and safety information will be collected as appropriate.

All the trial drugs and protocol-required laboratory investigations are provided free of charge to participants in both the arms throughout the trial.

### Outcomes

The primary effectiveness outcome is the difference in change in home seated mean SBP from randomization to month 6. The primary safety outcome is the percentage of participants who discontinue the drugs due to adverse events from randomization to month 6. Secondary outcomes include the proportion of participants achieving clinic BP control (<140/90 mmHg; <130/80 mmHg) and home BP targets (<135/85 mmHg and <130/80 mmHg) at Months 1, 3, and 6; changes in home and clinic DBP from randomization to follow-up visits; the incidence of adverse events of special interest (AESIs), including symptomatic hypotension, cough, abnormal laboratory findings, headache, peripheral oedema, and other events leading to permanent discontinuation of trial drug; adherence to BP-lowering drugs; modifications to BP-lowering therapy; and time to BP control.

### Sample size

Assuming a common standard deviation of home SBP of 12 mmHg, a sample size of 300 randomized participants (including 10% lost to follow-up) allocated in a 1:1 ratio to the two groups will provide >90% power at a two-sided significance level of α=0.05 to detect a difference in change in home SBP of at least 5 mmHg between groups. Clinic BP control (SBP <140 mmHg and DBP <90 mmHg) among treated patients in hospital settings in India has been estimated to be up to 50%. Assuming that 76% of participants in the intervention group and 60% in the control group achieve clinic BP control at Month 6, the sample size of 300 randomized participants will provide 80% power at a two-sided significance level of α=0.05 to detect a difference of at least 16% in clinic BP control at month 6.[21–23]

### Statistical analysis

All effectiveness analyses will be conducted according to the intention-to-treat principle and include all randomized participants. Baseline characteristics of randomised participants will be summarized by treatment group using appropriate descriptive statistics. The primary outcome, change in home systolic blood pressure (SBP) from randomization to month 6, will be analysed using a mixed-effects repeated-measures model (MMRM) including treatment group, visit, treatment-by-visit interaction, study site, baseline antihypertensive treatment status, age, sex, body mass index, and education level as fixed effects, with within-participant correlation accounted for through an appropriate covariance structure. Treatment effects will be presented as adjusted mean differences with 95% confidence intervals. A per-protocol (PP) analysis will also be done for the primary effectiveness outcome only. Secondary continuous BP outcomes will be analysed using similar MMRM approaches, while binary outcomes, including BP control rates, will be summarized as proportions and compared using adjusted log-binomial regression models to estimate risk ratios and risk differences with 95% confidence intervals. Safety analyses will be descriptive and performed in all participants who received at least one dose of study treatment, with adverse events of special interest and serious adverse events summarized by treatment group. Missing outcome data will be handled using the MMRM framework under the missing-at-random assumption. Exploratory subgroup analysis will be conducted by age, sex, BMI, home and clinic BP categories at randomisation, diabetes status and pre-randomisation drugs and these will be presented in a forest plot. More details of the analyses are published in a statistical analysis plan (Appendix 1).

### Current trial status

Between October 2025 and March 2026, 360 participants were screened and 300 were randomized from a total of nine sites (150 participants to the NewSTeP STP and 150 to the usual care group). A total of 60 participants were not randomized. Common reasons for screen failure included average BP

<135/85 mmHg, unwillingness to undertake protocol-specified home BP monitoring, and treatment with ≥2 BP-lowering medications. The last patient last visit for the randomized period is expected in September 2026, and the results of the randomized period are expected to be reported in October 2026.

### Baseline Characteristics

Baseline characteristics are presented in Table 2. The mean age of the participants at baseline was 51 years and 43% were women. The mean body mass index (BMI) was 28.2 kg/m^2^. Nearly half of participants had diabetes (49%), while dyslipidaemia (28%) and thyroid disorders (15%) were also common comorbidities. The sample included individuals with varied educational backgrounds; however, primary (33%) and secondary education (31%) were the most commonly reported levels of education. Smoking and alcohol use were relatively uncommon. The mean clinic BP at randomisation was 151/92 mmHg and mean home BP was 142/89 mmHg. About 51% were untreated/newly diagnosed at randomisation and 49% were taking a single antihypertensive drug.

**Table 2:** Baseline characteristics of included participants.

| **Participant characteristics** | **Overall (N = 300)** |
| --- | --- |
| **Demographics** |  |
| Age, years, mean (SD) | 51.4 (10.8) |
| Male sex, n (%) | 170 (56.7) |
| Female sex, n (%) | 130 (43.3) |
| **Education, n (%)** |  |
| No formal education | 18 (6) |
| Primary school ($\geq 6$ years) | 98 (32.7) |
| Secondary school ($\geq 12$ years) | 92 (30.7) |
| Tertiary education ($\geq 16$ years) | 70 (23.3) |
| Vocational training | 22 (7.3) |
| **Lifestyle factors** |  |
| BMI, kg/m<sup>2</sup>, mean (SD) | 28.2 (4.2) |
| Never smoked, n (%) | 283 (94.3) |
| Ex-smoker, n (%) | 4 (1.3) |
| Current smoker, n (%) | 13 (4.3) |
| Current alcohol use, n (%) | 34 (11.3) |
| **Medical history, n (%)** |  |
| Hypertension | 300 (100) |
| Diabetes | 147 (49) |
| Dyslipidaemia | 84 (28) |
| Thyroid disorder | 45 (15) |
| **Laboratory measures, mean (SD)** |  |
| Sodium, mmol/L | 138.4 (2.9) |
| Potassium, mmol/L | 4.3 (0.6) |
| Chloride, mmol/L | 101 (6.5) |
| Creatinine, mg/dL | 0.8 (0.2) |
| eGFR, mL/min/1.73 m<sup>2</sup> | 98.9 (20) |
| Blood glucose, mmol/L | 7.2 (3.4) |
| Total cholesterol, mmol/L | 4.9 (1.1) |
| LDL-C, mmol/L | 3.1 (0.9) |
| HDL-C, mmol/L | 1.2 (0.4) |
| Triglycerides, mmol/L | 2.0 (1.4) |
| **Antihypertensive drug at randomisation, n (%)** |  |
| None | 152 (50.7) |
| One drug | 148 (49.3) |
| **Blood pressure at randomisation, mean (SD), mmHg** |  |
| Clinic SBP/DBP | 151 (10.1)/ 92 (7) |
| Home SBP/ DBP | 142 (11.9)/ 89 (8.1) |

*Data are presented as mean (SD) for continuous variables and n (%) for categorical variables. BMI, body mass index; eGFR, estimated glomerular filtration rate; LDL-C, low-density lipoprotein cholesterol; HDL-C, high-density lipoprotein cholesterol; BP, blood pressure; SBP, systolic blood pressure; DBP, diastolic blood pressure.*

### Process evaluation

A mixed-methods process evaluation will be conducted alongside the trial to assess the implementation, acceptability, sustainability of the NewSTeP intervention. Guided by the RE-AIM (Reach, Effectiveness, Adoption, Implementation, and Maintenance) framework, the evaluation will combine quantitative trial data with qualitative findings to assess intervention delivery, explore mechanisms of impact, and identify factors influencing implementation across diverse healthcare settings. Quantitative measures will include recruitment and participation rates, BP control, drug use and adherence, quality of life, intervention fidelity, implementation costs, and indicators of sustainability.

To complement the quantitative evaluation, semi-structured interviews will be conducted with purposively selected patients, carers, and healthcare providers to capture a range of experiences and perspectives. Interviews will explore contextual influences on implementation, experiences with the treatment protocol, barriers and facilitators to adoption, acceptability of single-pill combination therapy, and perceptions of long-term sustainability. Interviews will be audio-recorded, transcribed verbatim, translated into English where required, and analysed using thematic analysis. Quantitative and qualitative findings will be integrated to provide a comprehensive understanding of intervention reach, effectiveness, adoption, implementation, and maintenance, and to inform the potential scale-up and integration of the intervention into routine hypertension care in India.

### Economic evaluation

A within-trial (6-month) and modelled lifetime economic evaluation will be conducted from the health system perspective to assess the cost-effectiveness of the intervention compared with usual care. Resource utilisation data, including healthcare visits, drug use, diagnostic tests, hospitalisations, and participant-incurred costs, will be collected during follow-up using healthcare diaries. The primary economic outcome will be the proportion of participants achieving BP control at 6 months, with cost-effectiveness expressed as the incremental cost-effectiveness ratio (ICER) per additional participant achieving BP control. A longer-term modelled analysis will estimate lifetime costs and disability-adjusted life years (DALYs) averted by extrapolating trial outcomes using a Markov model with health states including no cardiovascular disease, stroke, myocardial infarction, and death. Transition probabilities, costs, and disability weights will be derived from published evidence and local data sources. Lifetime cost-effectiveness will be expressed as the ICER per DALY averted, and sensitivity analyses will be undertaken to evaluate the robustness of findings and address uncertainty in key model parameters.

## Discussion

Hypertension remains the most important modifiable risk factor for cardiovascular disease, yet hypertension control rates in India remain low despite the availability of effective and affordable treatments. The NewSTeP trial was designed to address key gaps in hypertension management by evaluating a STP incorporating the early use of SPCs in routine clinical practice. This approach is supported by growing evidence that simplified treatment regimens and protocol-based care can improve BP control and reduce cardiovascular risk.

Despite guideline recommendations favouring early combination therapy, hypertension management in practice is still largely based on monotherapy, with treatment intensification often delayed. Such initial and subsequent therapeutic inertia contributes to prolonged periods of uncontrolled BP and increased cardiovascular risk. By combining SPC-based treatment with a structured intensification pathway, the NewSTeP intervention aims to address barriers such as treatment complexity, poor adherence, and variation in prescribing practices.[16, 24]

A key strength of the trial is its pragmatic design, which evaluates the intervention in routine healthcare settings, enhancing the generalisability and real-world applicability of the findings. The multicentre design and inclusion of diverse healthcare settings further strengthen the relevance of the findings for policy and practice. Alignment with the WHO HEARTS framework further enhances the relevance of the findings for hypertension control programmes in India and other low- and middle-income countries. The inclusion of process and economic evaluations is another key strength of the trial as it provides insights into implementation, scalability, and cost-effectiveness of the intervention alongside clinical outcomes.

A few limitations should be acknowledged. Firstly, the relatively small number of study sites may limit the representativeness of the participating settings and reduce generalisability to all healthcare contexts. The six-month follow-up period is adequate for assessing BP control and implementation outcomes but may not capture longer-term adherence or cardiovascular effects. In addition, the exclusion of patients with established cardiovascular disease, advanced kidney disease, severe hypertension, and other complex conditions may limit generalisability to higher-risk populations.

In conclusion, the NewSTeP trial will generate important evidence on the effectiveness and safety of a STP incorporating early SPC therapy and inform future hypertension control strategies in India and similar settings.

## Supporting information

Statistical Analysis Plan (Appendix 1)

## Ethics and consent

The trial protocol, participant information sheet and informed consent form were approved by the institutional ethics committee of each participating site before recruitment. The trial is being conducted in accordance with the Declaration of Helsinki, the principles of ICH Good Clinical Practice, and applicable ethical and Indian regulatory requirements. Written informed consent is obtained from every participant before any trial procedure.

## Funding

The NewSTeP trial is funded by a program grant from the Department of Biotechnology (DBT), Government of India and the Wellcome Trust, United Kingdom (India Alliance).

## Study Governance

### Trial conception and oversight

Abdul Salam, Vivekanand Jha, Anthony Rodgers, Mark D. Huffman, Dorairaj Prabhakaran, Stephen Jan

### Overall scientific oversight of the trial through the Steering Committee

Vivekanand Jha, Dorairaj Prabhakaran, Mark D. Huffman, Andrew Moran, Stephen Jan, Anthony Rodgers, Balram Bhargava, Rakesh Kumar Sahay, Gangadhar Taduri, Padinhare P. Mohanan, Sandeep Bansal, Prabhdeep Kaur, and Arpita Ghosh

### Site Principal Investigators

Sindhu Joshi, Rakesh Kumar Sahay, Viswanathan Mohan, Aneesh Basheer, Jayagopal Pathiyil Balagopalan, D.K.SriRam, Navneet Gill, Sanjay D Cruz, A. Venkateswara Rao

## Author contributions

### Trial implementation and project management

Abdul Salam, Krishnaiah Chappidi, Rupasvi Dhurjati, Rashmi Pant, Amit Kumar, Anshika Mittal, Shwetha Rajaram, Sasi Tiruttani, Gautam Satheesh, Parul Puri, Vivekanand Jha

### Manuscript writing

Rupasvi Dhurjati, Abdul Salam

### Critically reviewed the manuscript, provided intellectual input and approved the final version

All the authors

## Acknowledgements

The project team gratefully acknowledges Bijini Bahuleyan for developing and maintaining the REDCap-based electronic Case Report Forms (eCRFs), and the site staff for coordinating and overseeing project activities.

We also thank Anthony Rodgers, Balram Bhargava, Rakesh Kumar Sahay, Gangadhar Taduri, Padinhare P. Mohanan, Arpita Ghosh, Sanjay D Cruz for their invaluable support and contributions to the study.

## Competing Interests

MDH has received travel support from the World Heart Federation and consulting fees from Eli Lilly. MDH has pending patents for heart failure polypills. All other authors have no competing interest to declare.

## Patient and Public Involvement

Patients and the public were not involved in the design, conduct, reporting, or dissemination plans of this trial. The intervention and trial procedures were developed by the study investigators and expert committees, drawing on existing evidence and clinical and implementation experience in hypertension care.

## Data availability

Data are not currently available as the trial is ongoing. De-identified participant-level data underlying the final trial results may be made available upon reasonable request after publication of the main trial findings, subject to approval by the trial steering committee and relevant ethics and regulatory requirements.

## Supplementary file

Supplementary table 1: List of trial visits and assessments

## Appendix-1

Statistical Analysis Plan

## Notes

### Clinical Trial

CTRI/2025/07/091649

### Author Declarations

The study was approved by the institutional ethics committee of The George Institute for Global health and also each participating site before recruitment.

