## Supplementary file for "Effectiveness and safety of a New Standardised TrEatment Protocol based on initial/early therapy with single-pill combinations of blood pressure-lowering drugs for improving blood pressure control (NewSTeP): Rationale, design of a randomized clinical trial and baseline characteristics of the trial p"

**Short, abbreviated title (≤40 characters):** NewSTeP trial

**Corresponding Author**

Abdul Salam

The George Institute for Global Health

Shangrilla Plaza #401, Road No 2, Banjara Hills

Hyderabad, Telangana- 500034

**Supplementary table 1: List of trial visits and assessments**

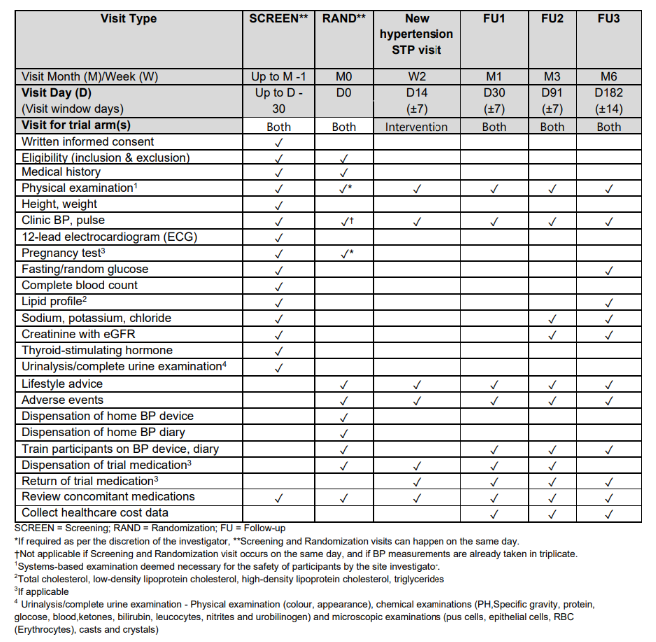
