## Supplementary material for "Effectiveness and safety of a New Standardised TrEatment Protocol based on initial/early therapy with single-pill combinations of blood pressure-lowering drugs for improving blood pressure control (NewSTeP): Rationale, design of a randomized clinical trial and baseline characteristics of the trial p": Statistical Analysis Plan (Appendix 1)

| | |
|---|---|
| Study Title | New Standardised TrEatment Protocol development and evaluation for improving blood pressure control |
| Brief Title | NewSTeP |
| Study/protocol Number | TGI-IA-HTN-AS-1 |
| Therapeutic Indication | Hypertension |
| Version and Date | Version 1.0 and date: 10/07/2026 |
| Study Registration | |
| Sponsor | The George Institute for Global Health, India |
| Funder | India Alliance - Department of Biotechnology (DBT), Government of India, and the Wellcome Trust, United Kingdom |

### TABLE OF CONTENTS

1 Administrative information 5

1.1 Revision history 5

1.2 Contributors to the statistical analysis plan 5

1.3 Approvals 5

2 Study synopsis 6

2.1 Study Objectives 6

2.2 Patient population 7

2.2.1 Inclusion criteria 7

2.2.2 Exclusion criteria 8

2.3 Outcomes 9

2.3.1 Primary Outcome 9

2.3.2 Secondary Outcomes 9

2.3.3 Primary Safety Outcomes 10

2.3.4 Tertiary/Exploratory Outcomes 10

2.4 Intervention 10

2.4.1 Intervention arm study treatment 10

2.4.2 Comparator arm study treatment 10

2.4.3 Referral to a specialist 11

2.4.4 Lifestyle advice 11

2.4.5 Down-titration or temporary discontinuation of the trial medication 11

2.4.6. Early Permanent Discontinuation of the Trial Medication 13

2.4.7. Add-On BP-lowering treatment 13

2.5 Randomisation and Blinding 13

2.6 Sample Size 14

2.7 Timing of outcome assessments 14

3 Statistical analysis 15

3.1 Computational environment 15

3.2 Interim analyses 15

3.3 Multi-center analysis 15

3.4 Multiplicity adjustment 16

3.5 Population for analysis 16

3.5.1 Intention to treat (ITT) 16

3.5.2 Per-protocol (PP) set 16

3.5.3 Safety set 17

3.6 Study disposition 17

3.7 Baseline characteristics 17

3.8 Adherence to study medications 18

### 1 Administrative information

#### 1.1 Revision history

This Statistical Analysis Plan (SAP) and all its subsequent versions (if any) will be prepared in accordance with guidelines proposed in the EQUATOR Network (1).

| Version | Date | Details |
|---|---|---|
| 1.0 | 13/01/2026 | Based on NewSTeP Study protocol Version 2.0 dated 27/05/2025 |

#### 1.2 Contributors to the statistical analysis plan

| Name | Affiliation | Responsibility |
|---|---|---|
| **Rashmi Pant** | The George Institute for Global Health, India | Principal Statistician will prepare the statistical analysis plan in consultation with the investigators, present blinded DSMB reports, blinded data monitoring and final data analysis post database unlock. |
| **Abdul Salam** | The George Institute for Global Health, India | Investigator will inform and review the Statistical analysis plan in accordance with the study objectives and the protocol. |
| **Parul Puri** | The George Institute for Global Health, India | Statistician (Unblinded) will generate the randomisation sequence, prepare unblinded DSMB reports, contribute to final data analysis. |

#### 1.3 Approvals

The undersigned have reviewed this plan and approved it as final. They find it to be consistent with the requirements of the protocol as it applies to their respective areas. They also find it to be compliant with ICH-E9 principles and, in particular, confirm that this analysis plan was developed in a completely blind manner (i.e. without knowledge of the effect of the intervention(s) being assessed).

| | | |
|---|---|---|
| Rashmi Pant | | 10/06/2026 |
| **Name** | **<signature>** | **<date>** |
| Abdul Salam | | 10/06/2026 |
| **Name** | **<signature>** | **<date>** |
| Parul Puri | | 10/06/2026 |
| **Name** | **<signature>** | **<date>** |

### 2 Study synopsis

The NewSTeP study involves a parallel arm randomised controlled trial designed to evaluate, among adults with hypertension, the effectiveness and safety of a new hypertension STP involving initial/early use of (fixed-dose combination) FDC of BP-lowering drugs compared to usual care for improving blood pressure control.

#### 2.1 Study Objectives

1. To evaluate, among adults with hypertension, the effectiveness of new STP involving initial/early use of FDC of BP-lowering drugs compared to usual care for improving blood pressure control.
2. To evaluate, among adults with hypertension, the safety of new STP involving initial/early use of FDC of BP-lowering drugs compared to usual care for improving blood pressure control.

**Figure 1.** Randomised clinical trial schema

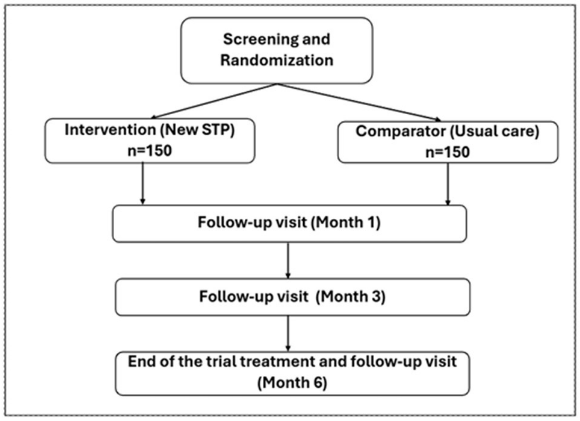

Note: For participants in the intervention arm, the additional follow-up visits will be as per the new hypertension STP, and for participants in the comparator arm, as per usual care

#### 2.2 Patient population

Participants will be recruited from the outpatient departments of healthcare centres (sites) providing hypertension care that have demonstrated capacity (i.e., the ability to recruit patients with hypertension as per the recruitment plan and conduct the trial procedures), with preference given to sites that participated in Aim 1 formative primary research and taking into account the diversity in geographic location of the site and patient characteristics.

##### 2.2.1 Inclusion criteria

Adults with hypertension, who are untreated or treated with one BP-lowering drug, and who could appropriately be treated with the new STP or usual care for hypertension.

**At screening visit**

1. Adults (age 18-79 years)
2. Have diagnosis of hypertension
    a. Documented previous diagnosis of hypertension and/or currently using BP-lowering drugs for hypertension
    b. Clinic attended automated seated mean SBP ≥140 mmHg and/or DBP ≥90 mmHg on screening visit day

       and

       At least one documented instance of ≥140 mmHg and/or DBP ≥90 mmHg in the past
3. Untreated for hypertension or on one BP-lowering drug for ≥2 weeks.

4. Needs initiation or intensification of BP-lowering medication(s) as per the investigator's judgement based on BP at this and/or previous visits
5. Willingness to use a home BP measurement device and measure BP at home for 6 months
6. Provided signed informed consent to participate in the trial

**At randomisation visit**

1. Untreated for hypertension or on one BP-lowering drug for ≥2 weeks
2. Clinic attended automated seated mean (average of last 2 measurements) SBP: 140-179 mmHg and/or DBP 90-109 mmHg
3. Willingness to use a home BP measurement device and measure BP at home for 6 months
4. Needs initiation or intensification of BP-lowering medication(s) as per the investigator's judgement based on BP at this and/or previous visits

##### 2.2.2 Exclusion criteria

**At screening visit**

1. Receiving 2 or more BP-lowering drugs (FDC containing two different BP-lowering drugs should be considered as 2 BP-lowering drugs)
2. Receiving any BP-lowering drugs for primary indications other than hypertension (e.g., migraine, benign prostate hyperplasia, heart failure)
3. Known current/history of secondary hypertension, cardiovascular disease, including coronary heart disease, angina, myocardial infarction, acute coronary syndrome, congestive heart failure, atrial fibrillation, stroke or transient ischemic attack
4. Known current/history of end-stage renal disease or anuria or current estimated glomerular filtration rate (eGFR) < 60 ml/min/1.73 $m^2$)
5. Women who are pregnant or had a positive pregnancy test or unwilling to take a pregnancy test before randomisation and/ or during the trial, breastfeeding, or of childbearing potential and not using effective contraception during the trial period.
6. Contraindication, including hypersensitivity (e.g., anaphylaxis or angioedema) to any of the trial medications or procedures
7. Participation in any investigational medication and/or device trial within the 30 days prior to randomisation

8. Current concomitant illness or physical impairment or mental condition or abnormal laboratory value, which in the judgment of the investigator could interfere with the effective conduct of the trial or constitutes a significant risk to the participants' safety or well-being

**At randomisation visit**

1. SBP ≥180 or DBP ≥110 mmHg
2. Fulfilling any of the exclusion criteria mentioned for the screening visit, when assessed again.

#### 2.3 Outcomes

The following are primary and secondary effectiveness and safety outcomes.

##### 2.3.1 Primary Effectiveness Outcome

Difference in change in home seated mean SBP from randomisation to month 6.

##### 2.3.2 Secondary Outcomes

1. Percentage of participants achieving clinic seated mean SBP <140 mmHg and DBP <90 mmHg at month 1, 3 and 6
2. Percentage of participants achieving home seated mean SBP <135 mmHg and DBP <85 mmHg at month 1, 3 and 6
3. Percentage of participants achieving clinic seated mean SBP <130 mmHg and DBP <80 mmHg at month 1, 3 and 6
4. Percentage of participants achieving home seated mean SBP <130 mmHg and DBP <80 mmHg at month 1, 3 and 6
5. Difference in change in home seated mean DBP from randomisation to 6 months
6. Difference in change in clinic seated mean SBP and DBP from randomisation to month 1, 3 and 6
7. Difference in change in home seated mean SBP and DBP from randomisation to month 1 and 3
8. Percentage of participants with incidence of adverse events of special interest (AESI) from randomisation to 6 months
9. Adherence to BP-lowering medications

10. Percentage of participants with modification of BP-lowering therapy, composite and separately (intensification, lessening, change, withdrawal) from randomisation to month 6
11. Time taken to achieve BP control

##### 2.3.3 Primary Safety Outcomes

Percentage of participants who discontinued trial medication due to AEs or SAEs from randomisation to month 6

##### 2.3.4 Tertiary/Exploratory Outcomes

1. Number of healthcare facility (including study site) visits related to hypertension
2. Percentage of participants treated with full adherence to the new hypertension STP
3. Percentage of referrals to hypertension specialists

#### 2.4 Intervention

Participants in both arms will be provided BP-lowering medications free of cost.

**Table 1.** Treatment arms in the trial

| Arms | Treatment arm | Participants | Treatment |
|---|---|---|---|
| 1 | Intervention | 150 | New hypertension standardised treatment protocol (STP) |
| 2 | Comparator | 150 | Usual care |

##### 2.4.1 Intervention arm study treatment

Participants in the intervention group will be treated following the new hypertension STP (Figure 2). Participants on one BP-lowering medication will be asked to switch to trial medications. Timing of the dosing of the trial medications will be as per the investigator's discretion.

Prescription of BP-lowering medications in addition to those required by new hypertension STP will be as per the discretion of the specialist and/or investigator.

##### 2.4.2 Comparator arm study treatment

Participants in the comparator arm will be treated as usual as per the discretion of the investigator.

##### 2.4.3 Referral to a specialist

In the intervention arm, participants with uncontrolled BP despite receiving the treatment following the new hypertension STP, will be referred to the specialist for evaluation and treatment advice. Specialists include hypertension specialist, cardiologists or nephrologists available at the healthcare centre in which the study site is located.

##### 2.4.4 Lifestyle advice

Participants in both the arms will be provided with the following standard lifestyle advice (2,3):

1. If using tobacco, stop use, and avoid harmful use of alcohol.
2. Engage in regular physical activity for at least 2.5 hours per week.
3. If overweight, lose weight.
4. Eat a heart-healthy diet low in salt, trans fats, and added sugar.

##### 2.4.5 Down-titration or temporary discontinuation of the trial medication

If a participant develops a condition or symptom likely to be related to the trial medication (e.g., due to possible hypotension) that is severe enough, the investigator can choose to down-titrate or discontinue the trial medication temporarily. The reason for down-titration or discontinuation will be recorded. If discontinued, then consideration should also be given to restarting trial medication subsequently, if benefits of restarting outweigh the risks as per the medical condition of the participant. If a participant discontinues medication at his/her own initiative, then the reason should be investigated and recorded. If medically appropriate, consideration should be given to restarting trial medication.

**Figure 2.** New hypertension standardised treatment protocol

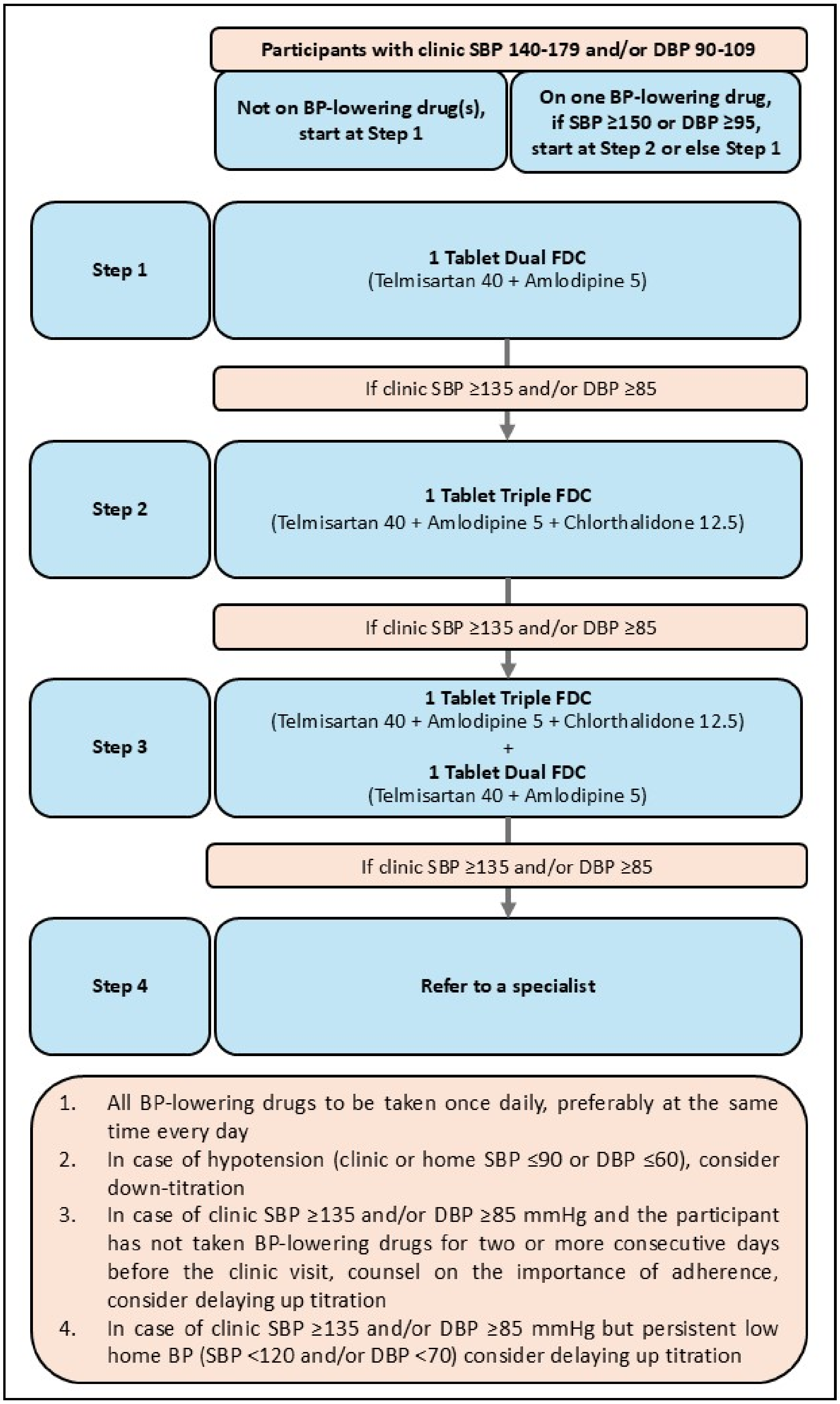
Participants with clinic SBP 140-179 and/or DBP 90-109
Not on BP-lowering drug(s), start at Step 1
On one BP-lowering drug, if SBP ≥150 or DBP ≥95, start at Step 2 or else Step 1
Step 1
1 Tablet Dual FDC
(Telmisartan 40 + Amlodipine 5)
If clinic SBP ≥135 and/or DBP ≥85
Step 2
1 Tablet Triple FDC
(Telmisartan 40 + Amlodipine 5 + Chlorthalidone 12.5)
If clinic SBP ≥135 and/or DBP ≥85
Step 3
1 Tablet Triple FDC
(Telmisartan 40 + Amlodipine 5 + Chlorthalidone 12.5)
+
1 Tablet Dual FDC
(Telmisartan 40 + Amlodipine 5)
If clinic SBP ≥135 and/or DBP ≥85
Step 4
Refer to a specialist
1. All BP-lowering drugs to be taken once daily, preferably at the same time every day
2. In case of hypotension (clinic or home SBP ≤90 or DBP ≤60), consider down-titration
3. In case of clinic SBP ≥135 and/or DBP ≥85 mmHg and the participant has not taken BP-lowering drugs for two or more consecutive days before the clinic visit, counsel on the importance of adherence, consider delaying up titration
4. In case of clinic SBP ≥135 and/or DBP ≥85 mmHg but persistent low home BP (SBP <120 and/or DBP <70) consider delaying up titration

##### 2.4.6. Early Permanent Discontinuation of the Trial Medication

Early permanent discontinuation of the trial medication occurs when a participant permanently ceases taking the trial medication before the end-of-trial treatment follow-up visit. Early permanent discontinuation of trial medication may happen for any of the following reasons:

1. By the participant themselves for any reason.
2. By the investigator at the request of the participant for any reason.
3. By the investigator based on clinical judgment or if participant's safety or well-being is or will be compromised by continued trial medication.
4. The trial is terminated.

Early permanent discontinuation should be recorded immediately along with the reason(s). Participants with early permanent discontinuation of the trial medication should be asked to continue their participation in the trial unaltered and such participants will still be followed up and all trial assessments performed as per the protocol until the end of the trial unless the participant withdraws consent.

##### 2.4.7. Add-On BP-lowering treatment

In intervention arm participants with uncontrolled BP despite receiving the treatment with STP, additional BP-lowering drugs can be provided to achieve blood pressure control. In the comparator, arm treatment will be as usual as per the discretion of the investigator and/or a specialist.

#### 2.5 Randomisation and Blinding

Randomisation sequence (1:1 allocation ratio) was generated using the R blockrand package with variable block sizes (2 and 4), stratified by site and baseline blood pressure medication status (untreated vs. treated). Each site was assigned a unique REDCap Data Access Group (DAG) code (e.g., Site 1 = 6406, Site 2 = 6407, ..., Site 15 = 6420), and participants were further stratified based on whether they were taking any BP-lowering medications before randomisation.

The randomisation list was created by a statistician not involved in day-to-day trial operations and uploaded to the REDCap randomisation module. Allocation concealment was maintained until the point of randomisation, which was triggered by site staff only after confirming participant eligibility and completing baseline assessments. The REDCap system then assigned the next available allocation from the appropriate stratum.

The trial is open label; participants and site investigators are aware of treatment allocation due to the nature of the interventions. However, outcome assessors and statisticians conducting the primary analysis will remain blinded to treatment allocation until database lock and finalisation of analysis code. Treatment groups will be coded as Arm A and Arm B during analysis to preserve blinding.

Personnel unblinded during trial conduct include site staff responsible for treatment implementation, the trial manager, REDCap administrator, and the statistician who generated the allocation table. The unblinded statistician will prepare confidential unblinded reports for DSMB review, including safety and effectiveness data by treatment group. All other trial personnel, including the blinded statistician and investigators, will remain unaware of treatment group outcomes until database lock.

#### 2.6 Sample Size

A sample size of 300 randomised participants (including 10% lost to follow-up) allocated in a 1:1 ratio to the two arms will provide >90% power at a two-sided significance level α=0.05 to detect a difference of at least 5 mmHg in home SBP (assuming a common standard deviation of 12 mmHg in both groups). Clinic BP control (SBP<140 and DBP <90 mmHg) in India, in hospital settings can be expected to be up to 50% (4). Assuming that at month 6, 76% of the participants will achieve clinic BP control in the intervention arm and 60% in the control arm (allowing for a better than normal result due to participation in the trial), the sample size of 300 randomised participants will also provide 80% power at a two-sided significance level α=0.05 to detect ≥16% difference in clinic BP control at month 6.

#### 2.7 Timing of outcome assessments

| **Visit Type** | **SCREEN**** | **RAND**** | **New hypertension STP visit** | **FU1** | **FU2** | **FU3** |
|---|---|---|---|---|---|---|
| Visit Month (M)/Week (W) | Up to M -1 | M0 | W2 | M1 | M3 | M6 |
| **Visit Day (D)**<br>(Visit window days) | Up to D -30 | D0 | D14<br>(±7) | D30<br>(±7) | D91<br>(±7) | D182<br>(±14) |
| **Visit for trial arm(s)** | Both | Both | Intervention | Both | Both | Both |

| | | | | | | |
|---|---|---|---|---|---|---|
| Physical examination | ✓ | ✓* | ✓ | ✓ | ✓ | ✓ |
| Height, weight | ✓ | | | | | |
| Clinic BP, pulse | ✓ | ✓† | ✓ | ✓ | ✓ | ✓ |
| 12-lead electrocardiogram | ✓ | | | | | |
| Fasting/random glucose | ✓ | | | | | ✓ |
| Complete blood count | ✓ | | | | | |
| Lipid profile | ✓ | | | | | ✓ |
| Sodium, potassium, chloride | ✓ | | | | ✓ | ✓ |
| Creatinine with eGFR | ✓ | | | | ✓ | ✓ |
| Thyroid-stimulating | ✓ | | | | | |
| Urinalysis/complete urine | ✓ | | | | | |
| Adverse events | | ✓ | ✓ | ✓ | ✓ | ✓ |
| Healthcare cost data | | | | ✓ | ✓ | ✓ |
| Trial medication | ✓ | ✓ | ✓ | ✓ | ✓ | |

### 3 Statistical analysis

#### 3.1 Computational environment

Analyses will be conducted primarily using STATA Version 19 (StataCorp. 2023. Stata Statistical Software: Release 19. College Station, TX: StataCorp LLC). R version 4.2.1 will also be used for visual representation.

#### 3.2 Interim analysis

The DSMB will meet at least twice throughout the duration of the trial for data review. Confidential unblinded reports prepared by the unblinded statistician. These reports will include unblinded safety data (adverse events, serious adverse events, and adverse events of special interest by treatment group) and unblinded effectiveness data (primary and key secondary outcomes by treatment group). All DSMB deliberations and unblinded data will remain confidential from the trial team, including the blinded statistician and investigators, until database lock. No stopping rules for effectiveness are pre-specified; the DSMB will advise the Trial Steering Committee if safety concerns warrant early termination.

#### 3.3 Multi-center analysis

Analysis of demographic and baseline characteristics, effectiveness and safety data will be pooled across all sites.

#### 3.4 Multiplicity adjustment

P-values will only be reported for the primary effectiveness outcome (home BP), key secondary effectiveness outcome (clinic BP) and primary safety outcome and no multiplicity adjustments will be conducted. All tests will be two-sided with a nominal α-level set at 0.05 (5%). Reporting secondary and exploratory end points will be limited to point estimates of effects with 95% confidence intervals. The widths of the intervals will not be adjusted for multiplicity, and the interpretation of these confidence intervals will avoid the language of definitive conclusions used to report statistically significant findings as assessed by formal hypothesis testing.

Incidence of adverse events of special interest (AESI) and Serious adverse events (SAEs) overall and by system organ class will not have p-values computed but rather will be presented descriptively. Other outcomes will not have p-values for treatment effect computed. Subgroup analyses will be conducted for the outcomes mentioned in section 3.10.3 with point estimates and 95% CIs for each subgroup category presented as forest plots and p-value for heterogeneity within each subgroup will be presented.

#### 3.5 Population for analysis

##### 3.5.1 Intention to treat (ITT)

The primary population for all planned analyses will be the randomised set, defined as all participants who met the eligibility criteria, provided written consent and were randomised to one of the two treatment groups. The randomised set will be the set of reference for analysing primary and secondary outcomes to preserve the intention-to-treat (ITT) principle.

##### 3.5.2 Per-protocol (PP) set

Per protocol analyses will be done for the primary effectiveness outcome only. Subjects will be excluded from the PP analysis if any of the following occur:

- Incorrectly included based on inclusion/exclusion criteria stated in section 2.2.
- Non-compliance: Adherence (as defined in section 3.8) to treatment protocol is <80%
- Wrong treatment: Received a treatment/intervention other than the one to which they were randomised.
- Missing primary outcome: No evaluable primary outcome data available after the baseline assessment.

##### 3.5.3 Safety set

For the analysis of adverse events, the safety set, defined as participants who took at least one dose of any one of the two study treatments protocols, will be used.

#### 3.6 Study disposition

All participants screened who met study inclusion criteria will be accounted for. The number of participants who were randomised; and reasons for ineligibility for randomisation will be summarised.

Participant disposition summaries will be based on the screened set and tabulated for the following categories:

- Number (%) of participants screened.
- Number (%) of participants randomised, ineligible for randomisation.
- Number (%) of participants randomised but did not receive study treatment.
- Number (%) of participants completing the study (defined as attending month 6 visit)
- Number (%) of participants prematurely discontinuing the study medications and reasons.
- Number (%) of participants prematurely discontinuing from the study and reasons.

#### 3.7 Baseline characteristics

Description of participant characteristics at randomisation (baseline) will be presented by treatment group. Discrete variables will be summarised by frequencies and percentages. Percentages will be calculated according to the number of participants for whom data are available. Continuous variables will be summarised using mean and Standard Deviation (SD), and median and interquartile limits (Q1-Q3), minimum and maximum values. Baseline measures for all randomised participants will be tabulated for the variables listed below.

- Demographic characteristics: age (years), sex, height (cm), weight (kg), BMI (kg/m2), education
- Medical history (health conditions, BP medications)

- Lifestyle (smoking, alcohol use)
- Randomisation (baseline) clinic BP is, BP measured in clinic with the clinic BP device on the day of randomisation.
- Randomisation (baseline) home BP is, BP measured at using home BP device after randomisation and before the start of study treatment with derivations defined in section 3.9.1.

For other variables (e.g., age, concomitant health conditions) baseline is, values from the screening or the randomisation visit. The screening and randomisation visits will either be on the same day or a maximum of 30 days apart.

#### 3.8 Adherence to study medications

Adherence will be evaluated using a pill count data reported for the periods in Month 1, Month 3, and Month 6. For each participant and for each period, the number of pills apparently taken will be obtained by subtracting the total number of pills returned and the total number of pills reported as lost from the total number of pills dispensed. The number of pills expected to be taken will be calculated by multiplying the number of expected treatment days during the relevant period by the daily dosing frequency that is recorded. Adherence will then be calculated as:

Adherence (%) = (pills dispensed - pills returned - pills lost) ÷ (expected treatment days × daily dosing frequency) × 100.

For treatment group 1 the Month 1 adherence period will include pill-accountability data from the Randomisation and Week 2 periods, while for treatment group 2 it will include data from Randomisation only. In the case of the Month 3 adherence period, the pill-accountability data taken at the Month 1 visit will be used, and for the Month 6 adherence period the data recorded at the Month 3 visit will be used. The visit stop date, or if the stop date is not available, the date of the next visit, will be counted as an expected pill-taking day. The raw adherence values will be kept for data-quality purposes, whereas the adherence percentages used in the summary analyses will be set at 100%. When pill-return data or any other information necessary for calculating adherence is missing, adherence for that participant-period will be stated as missing and will not be imputed. If the number of pills reported as lost is missing but the numbers dispensed and returned are available, the number lost will be assumed to be zero and this imputation will be noted. Adherence will be summarised at each clinic visit by giving the number of participants with observed adherence, the number with missing adherence, and the mean and distribution of the observed adherence. The number and percentage of participants who achieve adherence above 80% will also be given for each adherence period and treatment group..

#### 3.9 Data Availability and Definitions

During the trial, all BP measurements in clinic and at home will be made in seated position, therefore, any mention of BP measurements and values in this SAP implies BP at seated position.

The following study periods will be referenced for the main analysis using the randomisation set.

Day 0 = Day of randomisation

Day 1 = The day (00:00 to 23:59 hrs) after the day of randomisation.

Week 2, Month 1, 3, 6 follow-up clinic visits = Actual date of in-person or telephonic clinic visit (i.e., date of assessment in eCRF) after randomisation. Telephonic visits are conducted if the patient does not attend the scheduled clinic visit in person. Patients are asked to send screen shots of home BP measurements recorded in the diary via WhatsApp. These will be the reference dates for the calculation of home BP averages for each visit. **NB** these visits may not have occurred at these calendar timepoints because of various administrative or patient-related factors. A visit window will be applied to assign data to the correct scheduled visit: data collected within ±7 days of the scheduled Month 1 and Month 3 visit dates, and within ±14 days of the scheduled Month 6 visit date will be attributed to that visit. Where a visit falls outside these windows, data will be assigned to the nearest scheduled visit timepoint. Nonetheless, all data will still be ascribed to a visit name and included in the analysis regardless of whether the visit occurred within the protocol-specified window. Month 1, 3 and 6 calendar period = 30 (±7), 91 (±7), and 182 (±14) days since day of randomisation. These will be the reference periods for calculating BP means from all available BP values recorded since randomisation.

Week is defined in relation to the randomisation date with Week 1 beginning on Day 1, the first treatment with randomised trial medication: $Week = (date\ of\ BP\ measurement - date\ of\ first\ dose\ of\ randomized\ treatment)/7$

Blood pressure data availability will be summarised by follow-up visit periods (Week 2, Month 1, Month 3 and Month 6) and by treatment arm. For home BP, the following will be reported for each period: (i) the number and percentage of participants with at least one home BP measurement; (ii) the number and percentage with no available measurements; and (iii) the mean (SD) number of measurement per participant (iv) the mean (SD) number of triplicate measurements per participant (v) the number and percentage of participants who took BP measurements per protocol in the week prior to the scheduled clinic visit. For clinic BP, the number and percentage of participants with a valid clinic BP (average of the last two measurements in a triplicate set taken at least 1-minute intervals) reading at each scheduled visit will be reported separately.

##### 3.9.1 Description & Derivation of Home and Clinic BP

A valid home BP measurement is defined as one SBP (with the plausible range 81 to 249 mmHg) or DBP value (with the plausible range 41 to 149 mmHg), entered in the home BP diary and transcribed to the eCRF. Each participant is expected to record BP measurements in triplicate in a single sitting of 3–5-minute intervals. Details of BP measurement procedure are reported in the study protocol.

**Randomisation Home BP for analysis:** this will be mean of all valid home BP measurements on the day of randomisation (Day 0) and Day 1 (00:00 AM to 11:59 AM) (NB participants are asked to take home BP in the morning before taking study medication). If participants did not take-home BP measurements at home, we will use data from home BP device used in the clinic on the day of randomisation to calculate home BP at baseline.

**Follow up Home BP for a given month:** This will be mean of all valid home BP measurements taken between follow-up clinic visits including measurements taken on the morning of the given monthly clinic visit. For example, for month 1 follow up BP, if month 1 clinic visit date is 31 May, average of all valid home BP measurements on or between day 2 from randomisation and 31st May.

SBP and DBP measurements will be dealt with independently, that is, if SBP value is missing or outside the plausible range this will have no bearing on DBP estimation and vice versa.

Clinic BP at a scheduled monthly visit will be the mean of the last two readings of the triplicate as measured by the health facility staff during the clinic visit. Mean will be pre-calculated in the eCRF.

##### 3.9.2 Definitions & Derivation of adverse events and adverse events of special interest

An adverse event (AE) is any untoward medical occurrence (including a symptom / disease or an abnormal laboratory finding) during treatment with a pharmaceutical product in patient or a human volunteer that does not necessarily have a relationship with the treatment being given. For this trial, data will be collected only for AEs that are defined as AESI (Adverse Events of Special Interest).

***Adverse Events of Special Interest.*** The following AEs are considered AESI, and data will be collected for them:

Symptomatic hypotension: Dizziness or any other symptom or event possibly related to hypotension.

Cough

Abnormal laboratory findings of serum sodium, serum potassium, serum uric acid, serum creatinine, or eGFR.

Headache, including number of days with headache in the last month.

Peripheral oedema.

Any other symptoms or laboratory abnormality that led to permanent discontinuation of trial medication.

***Serious Adverse Events.*** An AE or adverse drug reaction that is associated with death, inpatient hospitalisation (in case the trial was being conducted on out-patients), prolongation of hospitalisation (in case the trial was being conducted on in-patients), persistent or significant disability or incapacity, a congenital anomaly or birth defect, or is otherwise life threatening.

***Intensity/Severity of an AESI/SAE.*** All AESI/SAEs will be graded as mild, moderate, or severe by the investigator based on her/his medical judgment and the following guidance:

Mild: asymptomatic or mild symptoms; clinical or diagnostic observations only; intervention not indicated.

Moderate: limiting age-appropriate instrumental activities of daily living (e.g. preparing meals, shopping for groceries or clothes, using the telephone); minimal, local, or non-invasive intervention indicated.

Severe: Medically significant but not immediately life-threatening; disabling; limiting self-care activities of daily living (e.g. bathing, dressing and undressing, feeding self); hospitalisation or prolongation of hospitalisation indicated.

***Relationship to the trial Medication.*** All AESIs/SAEs will be assessed for causal relationship to the trial medication by the investigator, and reported as definitely, probably, possibly, unlikely related, or not related.

***Suspected Unexpected Serious Adverse Reaction.*** A suspected unexpected serious adverse reaction (SUSAR) is a suspected adverse reaction related to an investigational medicinal product that is both unexpected and serious. An SAE will be considered unexpected if the nature, severity, or frequency of the event is not consistent with the information previously described for the triple pill in the Investigator Brochure (IB), or Prescribing Information.

#### 3.10 Analysis of the primary outcome

The primary effectiveness outcome of interest is the difference in home SBP change from randomisation to follow up visit month 6 between the intervention and comparator. This is estimated using statistical models with BP change from randomisation to follow up as dependent variable. For each participant, BP change is calculated as the BP at follow up minus the BP at randomisation. The estimate of the effectiveness of outcome will be made using directly measured assuming outcome data to be at least missing at random (MAR), in keeping with ITT analysis principles.

##### 3.10.1 Main analysis

Observed actual BP values, recorded by patients, will be summarised (see section 3.9.1) by follow-up visit timepoints and intervention and comparator for the primary effectiveness models. Mean BP at each clinic visit timepoint will be plotted (mean with 95% confidence interval plots) months since randomisation (for home and clinic BP).

The observed change in BP from randomisation to month 1, 3 and 6, for each participant, will be the difference between home BP at 1, 3 and 6-month follow up and home BP at randomisation as defined in section 3.9. This will be summarised by clinic visit timepoint and intervention group using means and 95% confidence intervals.

The modelled (estimated) difference in BP change from baseline (called mean BP reduction) will be based on the following approach (5) for ITT-analyses that provide valid inferences with missing data when outcome data are assumed at least MAR:

Mixed effect repeated measures model (MMRM) will be the primary effectiveness analysis model. The dependent variable will be the home BP change from randomisation to month 1, 3 and month 6 with fixed effects for treatment group, time (visit as categorical variable), treatment by time (visit) interaction, site, prior BP medications at randomisation (naïve or on monotherapy, age, sex, BMI and education (dichotomised as no schooling vs. some schooling). The variance will be estimated using a Huber-White sandwich estimator and the model will account for correlation within participant. An unstructured covariance matrix for participant observations will be used with fallbacks to first a compound symmetry (exchangeable) matrix and finally an independent matrix if the model does not converge. A compound symmetry covariance matrix will account for correlation within site with a fall back of an independent matrix if the model does not converge. Mean differences (Least square mean) and corresponding standard errors will be estimated for each follow-up visit and treatment group.

The following comparison will be extracted from the models as the estimated primary effectiveness endpoint.

Mean difference in SBP change from baseline (SBP reduction) at month 6, new treatment protocol vs usual care protocol.

Superiority of the intervention over the usual care will be shown when the comparison listed above will be statistically significant (when the null hypothesis of equal adjusted means is rejected). Mean difference in BP change at month 1, and 3 will also be extracted and p-values used only if superiority is observed at month 6.

##### 3.10.2 Adjusted analysis

The primary effectiveness analysis model will be adjusted for covariates described in section 3.10.1.

##### 3.10.3 Subgroup analyses

Subgroup analyses will be conducted for the primary effectiveness outcome-home SBP, home DBP and two selected additional safety outcomes (proportion with any AESI; and proportion with symptoms of hypotension). Subgroups for safety outcomes will only be conducted if more than 25 events are observed overall. Pre-specified subgroups are defined as follows:

- Sex (Male; Female)
- Age (≤50; >50 years)
- BMI (<30; >=30 kg/m$^2$),
- Diabetic; non-diabetic
- Randomisation clinic SBP category (<140, 140-159; ≥160 mmHg)
- Randomisation home SBP category (<140; 140-159, ≥160 mmHg)
- Pre-randomisation BP-lowering medications (0 or 1)

For each subgroup category, summary measures will include estimated mean differences with a 95% confidence interval for treatment vs control. These will be displayed using forest plots with p-values for difference in categories.

##### 3.10.4 Treatment of missing data

All reasonable efforts will be made to minimize missing data. The extent of missing data and reasons for missingness will be summarized by treatment group. Participant disposition, including missing primary outcome assessments and reasons for discontinuation or withdrawal, will be presented in the CONSORT flow diagram and in supporting listings.
The primary analysis will use a mixed model for repeated measures (MMRM), which uses all available observed outcome data without formal imputation. Under the assumption that data

are Missing At Random (MAR), the MMRM provides valid estimates of treatment effects. No ad hoc imputation methods (e.g., LOCF) will be used for the primary analysis.
If the proportion of missing primary outcome data is considered important, sensitivity analyses will be conducted to assess the robustness of conclusions to alternative assumptions about the missing data mechanism (section 3.10.5).

##### 3.10.5 Sensitivity analyses

The following sensitivity analyses will be conducted for primary effectiveness outcome only:

1. MMRM model for primary effectiveness outcome will be run using follow-up home BP at a given clinic visit month defined as mean of all valid home BP measurements taken 7-days prior and on the follow-up, clinic visit date.
2. An optional, reviewer requested, tipping point sensitivity analysis will be conducted (since this is not regulatory trial) only if the primary analysis demonstrates superiority of the treatment group at the 6-month follow-up. This will be done to test the consequences of the violation of the MAR assumption of the primary effectiveness analysis using the MMRM model described in section 3.10.1 and to assess the robustness of the primary effectiveness analysis under the Missing Not at Random (MNAR) assumption. The tipping point analysis will use a delta-adjusted multiple imputation approach. Missing values in the active treatment group will be imputed under the MAR assumption, and then systematically penalized by a pre-specified shift parameter $\delta$. The shift parameter will range from 0 to $\Delta$, exploring scenarios where the unobserved data are progressively worse than the observed data. The analysis will identify the specific value of ($\delta$) where the treatment effect is no longer statistically significant (i.e., the tipping point). The clinical plausibility of this tipping point will be evaluated to determine the robustness of the trial conclusions.

##### 3.10.6 Protocol deviations

All site-specific protocol deviations will be reviewed and, if necessary, site corrective actions will be implemented to mitigate future deviations. Major protocol deviations as defined in the Protocol Deviation Plan (Appendix 3), will be aggregated and presented as appropriate at time of final study analysis. The Data Safety Monitoring Board meeting held prior to database lock will determine which protocol deviations are major; the major deviations will be removed from the PP population. This summary will include the number and percent of ITT and PP subjects overall with each deviation type within each randomized treatment group.

#### 3.11 Analysis of secondary outcomes

Binary outcomes such as BP control (home BP <135/85 mmHg, home BP<130/80 mmHg and clinic BP <140/90 mmHg, clinic BP<130/80 mmHg) will be described as percentages by intervention group for each clinic visit month. Differences in BP control between new SPC protocol and usual care will be estimated for month 6 clinic visit only, using risk ratio and risk difference and their 95% confidence intervals obtained using log-binomial regression models adjusted for baseline BP, pre-randomisation treatment, age, sex, BMI, education and treatment time interaction. P-values will be reported for models, but no multiplicity adjustment done.

Difference in change in clinic SBP/DBP and home DBP at month 1,3 and 6 will be estimated using MMRM process described in section 3.10.1. Estimates will be reported as mean BP reduction and 95% confidence intervals between SPC protocol and usual care. P-values will be reported but not adjusted for multiple testing.

Adherence to BP-lowering medications will be reported descriptively for the two intervention groups using five-point summaries (mean, standard deviation, median, minimum and maximum value).

Modification of BP-lowering therapy, composite and separately (intensification, lessening, change, withdrawal) will be summarised using counts and percentage for each intervention group by clinic visit timepoint.

Time taken to achieve BP control (home and clinic <130/80 mmHg separately) will be expressed in terms of Time in therapeutic range (TTR) and its plots. TTR expresses the percentage of BP measurements recorded within a given BP window. TTR for BP includes both the mean BP value and the degree of BP variability during the follow-up period for each individual, reflecting BP variation over time (within and outside of the target range). We will calculate the percentage of observations for the following SBP windows 110-130, 110-135 and 110-140 mmHg.

#### 3.12 Analysis of safety outcomes

##### 3.12.1 SAEs and AESIs

All safety analysis will be descriptive in nature and 95% confidence intervals for unadjusted risk will be presented. Number of events and numbers (%) of participants experiencing AESIs will be tabulated by treatment group received and overall cumulated at month 6 follow up.

AESI or SAEs that emerge or worsen on or after the beginning of study treatments will be summarised:

- Adverse events of Special Interest,
- Serious AE,
- Treatment Related AESIs,
- Treatment Related SAEs,
- AE leading to study medication discontinuation,
- AE leading to study discontinuation.

SAEs and drug related AESIs will also be tabulated separately by treatment group and severity and by relationship to study medication. In case of incomplete date, or missing information in regard to adverse event analysis, the worst-case scenario approach will be used.

##### 3.12.2 Laboratory data and vital signs

Blood hematology, biochemistry and urine parameters will be collected according to the schedule of events. Only scheduled visit results will be presented, and unscheduled visits will be discarded for the purpose of the analysis. The baseline value for each parameter will be the latest non missing value recorded before first intake of study medication (all values from screening to randomisation date included will be taken into account).

For consistency, estimated Glomerular Filtration Rate (GFR) will be rederived and estimated using the CKD-epi equation 2021:

$$eGFR_{cr} = 142 \text{ x } \min(S_{cr}/\kappa, 1)^{\alpha} \text{ x } \max(S_{cr}/\kappa, 1)^{-1.200} \text{ x } 0.9938^{Age} \text{ x } 1.012 \text{ [if female]}$$

where:

$S_{cr}$ = standardised serum creatinine in mg/dL

$\kappa$ = 0.7 (females) or 0.9 (males)

$\alpha$ = -0.241 (female) or -0.302 (male)

$\min(S_{cr}/\kappa, 1)$ is the minimum of $S_{cr}/\kappa$ or 1.0

$\max(S_{cr}/\kappa, 1)$ is the maximum of $S_{cr}/\kappa$ or 1.0

Age (years)

Actual values and changes from baseline will be descriptively summarised by treatment group. Well referenced clinical cut-off values will be used to create binary variables for some laboratory parameters.

The number and percentage of participants in each category (above/below cut-off) will be computed and summarised descriptively.

#### 3.13 Analysis of tertiary/exploratory outcomes

Other exploratory outcomes include, number of healthcare facilities (including study site) visits related to hypertension, participants treated with full adherence to the new treatment protocol and referrals to hypertension specialists. Each of these outcomes will be described as counts and percentages.

#### 3.14 Other analyses

There is scope for potential descriptive analyses such as individual patient trajectories based on age, sex, baseline blood pressure and pre-randomisation medication.

### Appendix 1. List of abbreviations

AE – Adverse Event

AESI – Adverse Event of Special Interest

ACEI – Angiotensin-Converting Enzyme Inhibitor

ARB – Angiotensin II Receptor Blocker

BB – Beta Blocker

BMI – Body Mass Index

BP – Blood Pressure

CCB – Calcium Channel Blocker

DBP – Diastolic Blood Pressure

DIU – Diuretic

EDC – Electronic Data Capture

eCRF – Electronic Case Report Form

ECG – Electrocardiogram

eGFR – Estimated Glomerular Filtration Rate

FDC – Fixed-Dose Combination

HTN – Hypertension

NewSTeP – New Standardised TrEatment Protocol

RBC – Red Blood Cells

RCT – Randomised Clinical Trial

SAE – Serious Adverse Event

SUSAR – Suspected Unexpected Serious Adverse Reaction

SBP – Systolic Blood Pressure

SPC – Single Pill Combination

STP – Standardised Treatment Protocol

### Appendix 2. Tables, Figures and Listings

#### List of Tables

**Table 1.** Study disposition- Screened set

| | Overall |
|---|---|
| **Pre-screened** | |
| **Eligible** | |
| **Eligible for screening** | |
| **Screened** | |
| **Ineligible for randomisation (1) *** | |
| **Randomised (1) **** | |
| **Completed the study (2) (3)** | |
| **Discontinued from the study (2)** | |
| **Reasons for discontinuation (4)** | |
| Withdrew consent | |
| Loss-to-follow up | |
| SAE | |

*Reasons for ineligibility needs to be mentioned for individual participant

*Note:*

*(1) the denominator is the number of participants who were screened.*

*(2) the denominator is the number of participants who were randomised.*

*(3) include all participants who were randomised and completed all follow-up visit independent of premature stopping of study medication.*

*(4) the denominator is the number of participants who discontinued from the study*

**Table 2.** Demographics details by treatment group - randomised set

| Participant Characteristics | New STP (N=150) | Usual Care (N=150) | Overall (N=300) |
|---|---|---|---|
| **Age (years)** | | | |
| N | | | |
| Missing | | | |
| Mean (SD) | | | |
| Median (Q1; Q3) | | | |
| Min, Max | | | |
| **Sex** | | | |
| N | | | |
| Missing | | | |
| Female | | | |
| Male | | | |
| **Education** | | | |
| N | | | |
| Missing | | | |
| No formal education | | | |
| Primary School | | | |
| Secondary School | | | |
| Tertiary Education | | | |
| Vocational Training | | | |
| **BMI (Kg/m$^2$)** | | | |
| N | | | |
| Missing | | | |
| Mean (SD) | | | |
| Median (Q1; Q3) | | | |
| Min, Max | | | |
| **Smoking** | | | |
| Never | | | |
| Former | | | |
| Current | | | |
| **Current Alcohol Consumption** | | | |
| Never | | | |
| Former | | | |
| Current | | | |
| **Medical history** | | | |
| Chronic kidney disease | | | |
| Diabetes | | | |
| Dyslipidaemia | | | |
| **Laboratory measures, mean (SD)** | | | |
| Sodium, mmol/L | | | |
| Potassium, mmol/L | | | |

| |
|---|
| Chloride, mmol/L |
| Calcium, mmol/L |
| Uric acid, mmol/L |
| Creatinine, mg/dL |
| eGFR, mL/min/1.73 m² |
| Spot urine albumin to creatinine ratio, mg/g |
| Blood glucose, mmol/L |
| Glycated hemoglobin, % |
| Total cholesterol, mmol/L |
| LDL-C, mmol/L |
| HDL-C, mmol/L |
| **BP lowering medication at randomisation** |
| 0 |
| 1 |
| **Clinic blood pressure at randomisation, mean (SD), mm Hg** |
| Systolic |
| Diastolic |
| **Clinic heart rate, mean (SD)** |
| **Home blood pressure at randomisation, mean (SD), mm Hg** |
| Systolic |
| Diastolic |

**Table 3.** Home BP summary by visit period

| | Home BP (mmHg) | | | | | |
|---|---|---|---|---|---|---|
| | SBP | | | DBP | | |
| | New STP | Usual Care | New STP | Usual Care | New STP | Usual Care |
| **Screening** | | | | | | |
| N (Participants) | | | | | | |
| Mean (SD) | | | | | | |
| Min, Max | | | | | | |
| **Randomisation** | | | | | | |
| N (Participants) | | | | | | |
| Mean (SD) | | | | | | |
| Min, Max | | | | | | |

| |
|---|
| **Follow-up Month 1** |
| N (Participants)<br>Mean (SD)<br>Min, Max |
| **Follow-up Month 3** |
| N (Participants)<br>Mean (SD)<br>Min, Max |
| **Follow-up Month 6** |
| N (Participants)<br>Mean (SD) |

**Table 4.** Clinic BP summary by visit period

| | SBP | | | DBP | | |
|---|---|---|---|---|---|---|
| | **New STP** | **Usual Care** | **Overall** | **New STP** | **Usual Care** | **Overall** |
| **Screening**<br>N (Participants)<br>Mean (SD)<br>Min, Max | | | | | | |
| **Randomisation** | | | | | | |
| N (Participants)<br>Mean (SD)<br>Min, Max | | | | | | |
| **Follow-up Month 1** | | | | | | |
| N (Participants)<br>Mean (SD)<br>Min, Max | | | | | | |
| **Follow-up Month 3** | | | | | | |
| N (Participants)<br>Mean (SD)<br>Min, Max | | | | | | |
| **Follow-up Month 6** | | | | | | |
| N (Participants)<br>Mean (SD)<br>Min, Max | | | | | | |

**Table 5A.** Blood Pressure control status for home thresholds

| | New STP | Usual Care | Overall |
|---|---|---|---|
| **Follow-up Month 1** | | | |
| Home BP < 135/85 mmHg<br>Home BP < 130/80 mmHg | | | |
| **Follow-up Month 3** | | | |
| Home BP < 135/85 mmHg<br>Home BP < 130/80 mmHg | | | |
| **Follow-up Month 6** | | | |
| Home BP < 135/85 mmHg<br>Home BP < 130/80 mmHg | | | |

**Table 5B.** Blood Pressure control status for clinic thresholds

| | New STP | Usual Care | Overall |
|---|---|---|---|
| **Follow-up Month 1** | | | |
| Clinic BP <140/90 mmHg<br>Clinic BP <130/80 mmHg | | | |
| **Follow-up Month 3** | | | |
| Clinic BP <140/90 mmHg<br>Clinic BP <130/80 mmHg | | | |
| **Follow-up Month 6**<br>Clinic BP <140/90 mmHg<br>Clinic BP <130/80 mmHg | | | |

**Table 6.** Primary effectiveness outcome estimated mean difference in home SBP

| Visit | New STP Change from Baseline | Usual Care Change from Baseline | Treatment Mean Difference (95% CI) |
|---|---|---|---|
| Month 1 | | | |
| Month 3 | | | |
| Month 6 | | | |
| **Sensitivity Analysis-1** | | | |
| Month 1 | | | |
| Month 3 | | | |
| Month 6 | | | |
| **Sensitivity Analysis-2** | | | |
| Month 1 | | | |
| Month 3 | | | |

| |
|---|
| Month 6 |

**Note.** Sensitivity analysis 1 includes multiple imputation using multiple chained equations with covariates age, sex, education, BP at randomisation and BP medications at baseline; Sensitivity analysis 2 includes estimation of primary effectiveness outcome using home BP measurements taken 14 days prior to clinic visits during the randomisation phase

**Table 7.** Secondary effectiveness: changes in blood pressure

| Measure | New STP Change from Baseline | Usual Care Change from Baseline | Treatment Mean Difference (95% CI) | p-value |
|---|---|---|---|---|
| **Home DBP change from baseline** | | | | |
| Month 1 | | | | |
| Month 3 | | | | |
| Month 6 | | | | |
| **Clinic SBP change from baseline** | | | | |
| Month 1 | | | | |
| Month 3 | | | | |
| Month 6 | | | | |
| **Clinic DBP change from baseline** | | | | |
| Month 1 | | | | |
| Month 3 | | | | |
| Month 6 | | | | |

**Table 8.** Adherence to BP-lowering medications

| | New STP | Usual Care |
|---|---|---|
| **Month 1** | | |
| N | | |
| Mean (SD) | | |
| Median (Q1, Q3) | | |
| Min, Max | | |
| % <80% | | |
| % ≥80% and ≤120% | | |
| % >120% | | |
| **Month 3** | | |

| |
|---|
| N<br>Mean (SD)<br>Median (Q1, Q3)<br>Min, Max<br>% <80%<br>% ≥80% and ≤120%<br>% >120% |
| **Month 6** |
| N<br>Mean (SD)<br>Median (Q1, Q3)<br>Min, Max<br>% <80%<br>% ≥80% and ≤120%<br>% >120%<br>Overall |

**Note.** ≥ 80%: Ensures participants are taking at least 80% of the expected pills
≤120%: Caps adherence to avoid overconsumption

**Table 9**. Modification of BP-lowering therapy

| Visit | New STP | Usual Care | Overall |
|---|---|---|---|
| **Month 1** | | | |
| N<br>Eligible<br>Intensification (n, %)<br>Lessening (n, %) | | | |
| **Month 3** | | | |
| N<br>Eligible<br>Intensification (n, %)<br>Lessening (n, %) | | | |
| **Month 6** | | | |
| N<br>Eligible<br>Intensification (n, %)<br>Lessening (n, %) | | | |
| **Cumulative** | | | |
| N<br>Eligible<br>Intensification (n, %)<br>Lessening (n, %) | | | |

**Table 10.** Time in therapeutic range for Systolic Blood Pressure window

| SBP Window (mmHg) | New STP | Usual Care |
|---|---|---|
| **110-130** | | |
| Mean % (SD) | | |
| Median (Q1, Q3) | | |
| **110-135** | | |
| Mean % (SD) | | |
| Median (Q1, Q3) | | |
| **110-140** | | |
| Mean % (SD) | | |
| Median (Q1, Q3) | | |

**Table 11.** Safety: SAEs and AESIs

| Parameter | New STP | | Usual Care | | Overall | |
|---|---|---|---|---|---|---|
| | N | % | N | % | N | % |
| SAE | | | | | | |
| AESI | | | | | | |
| Treatment-related SAE | | | | | | |
| Treatment-related AESI | | | | | | |
| AE leading to medication discontinuation | | | | | | |

**Table 12.** Summary of Laboratory Parameters by Visit

| Parameter | Visit | Mean (SD) | | Overall – Mean (SD) | Min, Max (Overall) |
|---|---|---|---|---|---|
| | | New STP | Usual Care | | |
| **Potassium** | Baseline | | | | |
| | Month 1 | | | | |
| | Month 3 | | | | |
| | Month 6 | | | | |
| **Sodium** | Baseline | | | | |
| | Month 1 | | | | |
| | Month 3 | | | | |
| | Month 6 | | | | |
| **Creatinine** | Baseline | | | | |
| | Month 1 | | | | |
| | Month 3 | | | | |
| | Month 6 | | | | |
| **eGFR** | Baseline | | | | |
| | Month 1 | | | | |
| | Month 3 | | | | |
| | Month 6 | | | | |

**Table 13.** Subgroup analyses

| Sub-group | Treatment Difference (95% CI) | p-value |
|---|---|---|
| **Sex** | | |
| Male | | |
| Female | | |
| **Age** | | |
| ≤ 50 years | | |
| >50 years | | |
| **Body Mass Index (kg/m2)** | | |
| < 30 | | |
| ≥30 kg/m² | | |
| **Diabetes** | | |
| Yes | | |
| No | | |
| **Baseline clinic SBP category** | | |
| <140 mmHg | | |
| 140–159 mmHg | | |
| ≥160 mmHg | | |
| **Pre-randomised BP medications** | | |
| Naïve | | |
| Monotherapy | | |

#### List of Figures

Figure 1. New hypertension standardised treatment protocol (STP) diagram

Figure 2. CONSORT participant flow

Figure 3A. Home BP trajectories

Figure 3B. Clinic BP trajectories

Figure 4. Blood Pressure-Lowering Drug Regimens at each Follow-up visits

Figure 5. Scatter/Line Plot: primary & secondary treatment effects

Figure 6. Forest plot: sub-group analyses

Figure 7. Scatter Plot: Percentage of participants with >80% home BP measurements <130/80 versus follow-up BP at month 6

Figure 8. Time in therapeutic range (TTR), percentage of participants in therapeutic range at each time point

#### Listings

Listing 1. CONSORT listing: screened, randomised, analysed

Listing 2. Protocol deviations

Listing 3. SAEs details

Listing 4. AESIs details

Listing 5. Number of participants on different medication
